# Fear of High Blood Sugar in Pregnancy Questionnaire: development of a person-reported outcome measure to assess fear of hyperglycaemia in pregnant women with type 1 diabetes

**DOI:** 10.1101/2025.11.17.25340428

**Authors:** Jenny Gu, Amy Dunlop, Lois E Donovan, Kathleen H Chaput

**Author notes:** **Corresponding Author:** Lois E Donovan, Department of Medicine, Cumming School of Medicine, University of Calgary, Richmond Rd Diagnostic and Treatment Ctr, 1820 Richmond Rd SW, Calgary, Alberta T2T 5C7, Canada. Donovan and Chaput should be considered joint senior author. **Funding:** None. **Bulleted Novelty Statement:** -Diabetes management strategies that address medical and psychological concerns of those with type 1 diabetes during pregnancy are needed. Qualitative work has shown that technology improves diabetes management experiences in pregnancy. However, most studies show no quantitative benefits on person-reported outcome measures, likely because existing measures do not capture outcomes of importance to these individuals. We present a questionnaire to assess hyperglycaemia fear in pregnancy, an outcome that those living with type 1 diabetes have identified as important. Future research can begin to quantify the impact of new diabetes management strategies on hyperglycaemia fear in pregnancy.

## Abstract

**Background:** Although newer diabetes technologies improve glycaemic outcomes in type 1 diabetes pregnancies, most studies have not demonstrated improvements in scores on person-reported outcome measures (PROMs). This is likely because commonly used PROMs do not effectively capture quality-of-life concerns of greatest importance to pregnant women with type 1 diabetes.

**Aims:** We aimed to develop a research questionnaire to evaluate fear of hyperglycaemia in type 1 diabetes pregnancies, which is an important person-reported outcome in this population.

**Methods:** The format of the questionnaire was modelled after that of the Hypoglycaemia Fear Survey-II. Items pertaining to fear of hyperglycaemia, specifically in pregnancy, were developed. The questionnaire was refined using cognitive interviewing among Canadian women with experience of type 1 diabetes in pregnancy. We recruited participants sequentially and modified our questionnaire iteratively based on participants’ feedback. Saturation was reached when no significant changes were made by three consecutive participants.

**Results:** The questionnaire was reviewed by nine participants with median type 1 diabetes duration of 22 years. The questionnaire asks about 16 behaviours to avoid hyperglycaemia in pregnancy and 12 worries related to hyperglycaemia in pregnancy. 5-point Likert scales are used to quantify frequency of behaviours and worries.

**Conclusions:** Type 1 diabetes management is challenging in pregnancy. One source of stress for women is the fear of hyperglycaemia and its effects on their unborn children. The Fear of High Blood Sugar in Pregnancy Questionnaire is a pregnancy-specific person-reported outcome measure which can begin to quantify the impact of diabetes management strategies on hyperglycaemia fear.

## Introduction

During pregnancy, women with type 1 diabetes are advised to target blood glucose as close to the non-diabetes reference range as safely possible, in order to reduce the risk of maternal and neonatal complications.^1,2^ Despite this, consistently achieving target glucose levels for women with type 1 diabetes in pregnancy is rarely attained and attempts to do so can pose tremendous stress and increase diabetes management burden. Currently available insulin delivery methods and durations of exogenous insulins still fall short of mimicking the function of a healthy pancreas. As a result, pregnant women with type 1 diabetes remain at risk of severe hypoglycaemia and struggle to keep up with the changes in insulin requirements throughout pregnancy that are needed to reduce hyperglycaemia.^3^ Qualitative work by our group and others have highlighted the stress experienced by pregnant women with type 1 diabetes, related to fear of hyperglycaemia and possible consequences to their babies.^4-6^

Studies of diabetes technologies have shown improved time in target glucose range and reduced time above target glucose range in pregnancy with the use of continuous glucose monitoring^2^ and closed-loop insulin delivery.^7,8^ Qualitative interviews of women with type 1 diabetes in pregnancy in the Automated Insulin Delivery in Women with Pregnancy Complicated by Type 1 Diabetes (AiDAPT) study have highlighted that closed-loop insulin delivery can facilitate positive experiences of diabetes management in pregnancy.^5^ However, the AiDAPT study failed to demonstrate improvements on standardised diabetes person-reported outcome measures (PROMs).^7^ The CONCEPTT study similarly found that improved glycaemia with continuous glucose monitor use in pregnancy was accompanied by no improvements on standardised diabetes PROMs.^2^ We have shown that a group of women with type 1 diabetes felt that existing standardised questionnaires do not adequately capture the quality-of-life outcomes that are of greatest importance during pregnancy,^4^ which may explain the lack of observed benefits on PROMs and the discrepancy between the current qualitative and quantitative evidence surrounding type 1 diabetes in pregnancy.

Given the lack of a standardised PROM to address the expressed concern of hyperglycaemia in pregnancy among women with type 1 diabetes,^4^ we sought to develop a questionnaire to assess the fear of hyperglycaemia in pregnancy.

## Methods

### 1. Questionnaire development

The proposed Fear of High Blood Sugar in Pregnancy Questionnaire was designed to be self-administered by women with type 1 diabetes at any one time, or at multiple times, during pregnancy, for research purposes. The first iteration of the questionnaire was developed by a member of the research team with lived experience of type 1 diabetes in pregnancy and was modelled after the Hypoglycaemia Fear Survey-II (HFS-II), which is a well-validated tool to assess fear of hypoglycaemia in type 1 diabetes.^9,10^ The HFS-II asks about behaviours respondents engage in to avoid hypoglycaemia and worries related to lows. We developed a questionnaire with behaviour and worry subscales pertaining to hyperglycaemia and its consequences to both respondents and their babies. Item response options were 5-point Likert scales (“never”, “rarely”, “sometimes”, “often”, and “almost always”). To assess internal consistency of behaviour and worry subscales, we included questions about frequency of glucose monitoring, degree of hyperglycaemia-associated distress, and recent diabetic ketoacidosis.

### 2. Questionnaire revision through cognitive interviewing: study design

We conducted cognitive interviews with women with lived experience of type 1 diabetes and pregnancy to ensure that our questionnaire captured the range of behaviours and worries that women have related to fear of hyperglycaemia in pregnancy. Structured one-on-one interviews were conducted with participants and our questionnaire was modified iteratively based on each participant’s feedback as they reviewed the questionnaire. The Conjoint Health Research Ethics Board at the University of Calgary approved this study (REB 21-0447).

### 3. Study participants

We recruited Canadian participants through the University of Calgary’s *Participate-in-Research* website, and word-of-mouth messaging within the research team’s clinical networks. Individuals were eligible to participate in cognitive interviews if they were: 1) diagnosed with type 1 diabetes at least 12 months prior to a previous or current pregnancy; 2) able to speak and read English; 3) able to provide informed consent; and 4) able to meet with a member of the research team in person or had internet or phone access to meet virtually.

### 4. Study procedure

Participants completed a questionnaire that collected demographic, diabetes history, and pregnancy history data. The first three digits of participants’ postal codes were collected to determine neighbourhood median income.^11^

One-on-one interviews were conducted with each participant by one of two members of the research team. Participants were asked structured questions regarding each item in the behaviour and worry subscales of the questionnaire; specifically, whether each item was relevant, clear, focused, and non-judgmental. Participants were asked to propose alternative phrasings of items that were unsatisfactory, and to add new items that reflected experiences that were not already captured. The questionnaire was modified according to each participant’s responses, and the most updated version was used during each successive interview. If a participant proposed an edit that conflicted with that of a previous participant, both versions were presented to subsequent participants until a majority consensus was reached. We determined that saturation was reached when no significant changes were made to the questionnaire by three consecutive participants.

## Results

### 1. Participant characteristics

Our study included nine participants who ranged in age from 29 to 39 years. Participants were primarily white (88.9%) and had neighbourhood median annual household incomes over $80,000 Canadian dollars (77.8%) (Table 1). The median duration of diabetes was 22 years. Six out of nine participants were using an automated insulin delivery system at the time of study participation. Altogether the nine participants had experienced 25 pregnancies, three of which were ongoing at the time of study participation. Participants had children ranging in age from two months to 12 years.

**Table 1:** Participant characteristics.

|  |  |
| --- | --- |
| Age at enrolment in years (median (IQR)) | 36 (31-37) |
| Age at diabetes diagnosis in years (median (IQR)) | 10 (9-17) |
| Duration of diabetes in years (median (IQR)) | 22 (17-28) |
| Self-reported ethnic background (number (%)) |  |
| White/Caucasian | 8 (88.9) |
| African and Hispanic | 1 (11.1) |
| Maternal highest level of education completed (number (%)) |  |
| Secondary education | 1 (11.1) |
| University undergraduate degree or equivalent | 5 (55.6) |
| University higher degree or equivalent | 1 (11.1) |
| Trade or technical school degree or equivalent | 2 (22.2) |
| Neighbourhood median total income in Canadian dollars (number (%)) |  |
| \$70,000 to \$79,999 | 2 (22.2) |
| \$80,000 to \$89,999 | 2 (22.2) |
| \$110,000 to \$119,999 | 2 (22.2) |
| \$120,000 to \$129,999 | 2 (22.2) |
| \$140,000 to \$149,999 | 1 (11.1) |
| Number of pregnancies (number (%)) |  |
| 1 | 4 (44.4) |
| 2 | 1 (11.1) |
| 4 | 2 (22.2) |
| 5 | 1 (11.1) |
| 6 | 1 (11.1) |
| Pregnant at the time of study participation (number (%)) | 3 (33.3) |
| Number of children (number (%)) |  |
| 1 | 4 (44.4) |
| 2 | 4 (44.4) |
| 4 | 1 (11.1) |
| Age of children (median (IQR)) | 3.5 (2-5.5) |
| Current mode of insulin delivery (number (%)) |  |
| Standard insulin pump | 1 (11.1) |
| Automated insulin delivery (hybrid*) | 6 (66.7) |
| Multiple daily injections | 2 (22.2) |
| Diabetes complications (number (%)) |  |
| Yes | 2 (22.2) |
| No | 7 (77.8) |

Abbreviations: IQR, interquartile range.
\*Requires user to bolus insulin for meals

### 2. Questionnaire

Based on responses from the nine study participants, the proposed Fear of High Blood Sugar in Pregnancy Questionnaire was modified until the final version was developed (Table 2). In the behaviour subscale, participants suggested items be rephrased in order to clearly distinguish between personal/recreational and work-related contexts, since they felt that their behaviours to manage hyperglycaemia were dependent on the setting. Multiple items were added to the worry subscale based on participant feedback, including worries about perceived judgment from health care providers, inadequate diabetes support from health care providers, and hyperglycaemia affecting labour and delivery plans. The item, “Because my blood sugar could go high, I worry about my baby’s health” was moved to the beginning of the worry subscale due to one participant’s feedback that her baby’s health was the most important concern during pregnancy, above any concerns to her own health. One participant felt that it was judgmental to ask if respondents gave more insulin than the pump-recommended bolus to avoid hyperglycaemia, however acknowledged that there was not another way to phrase the item. The remainder of the items in the questionnaire were perceived as non-judgmental.

**Table 2:**
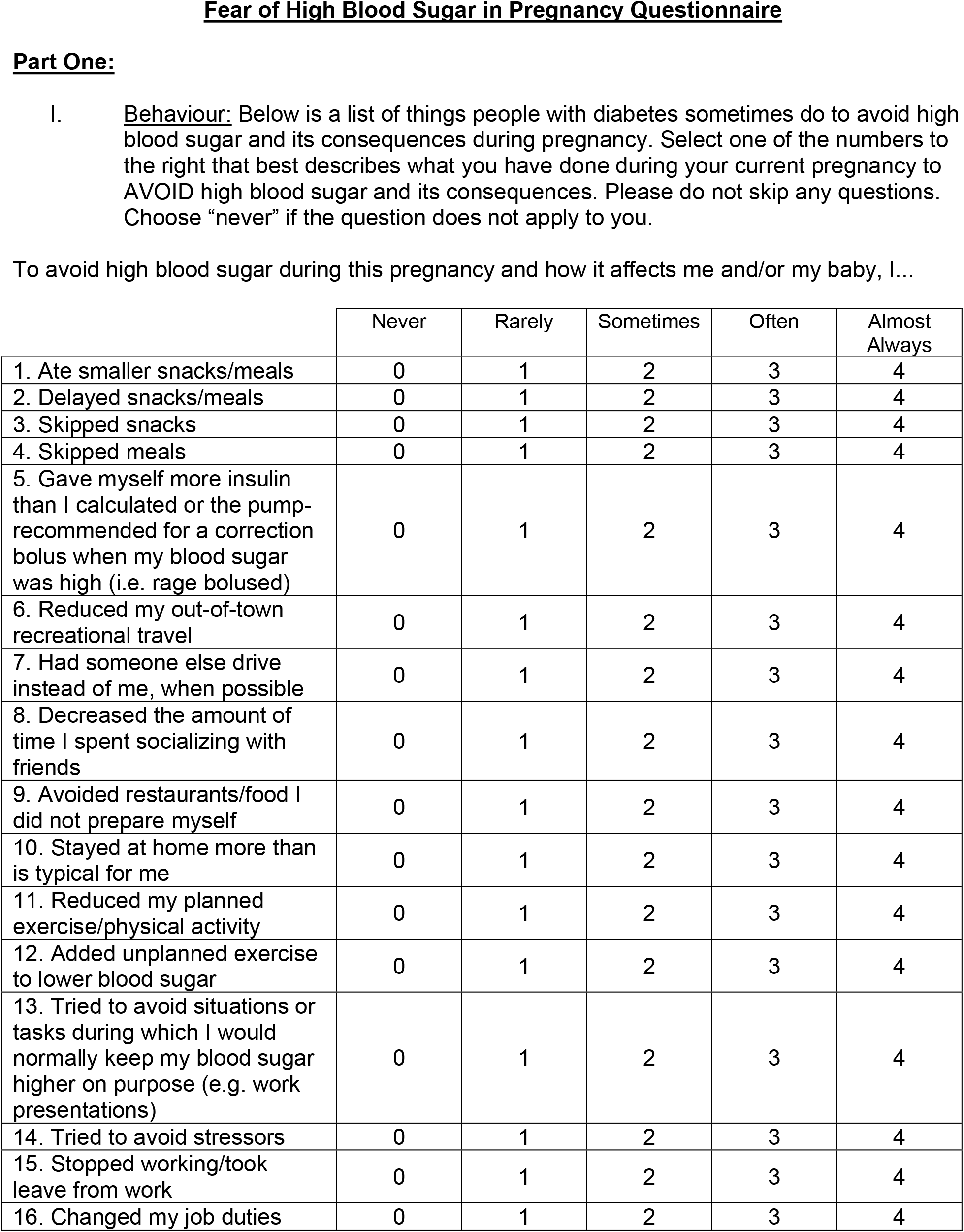
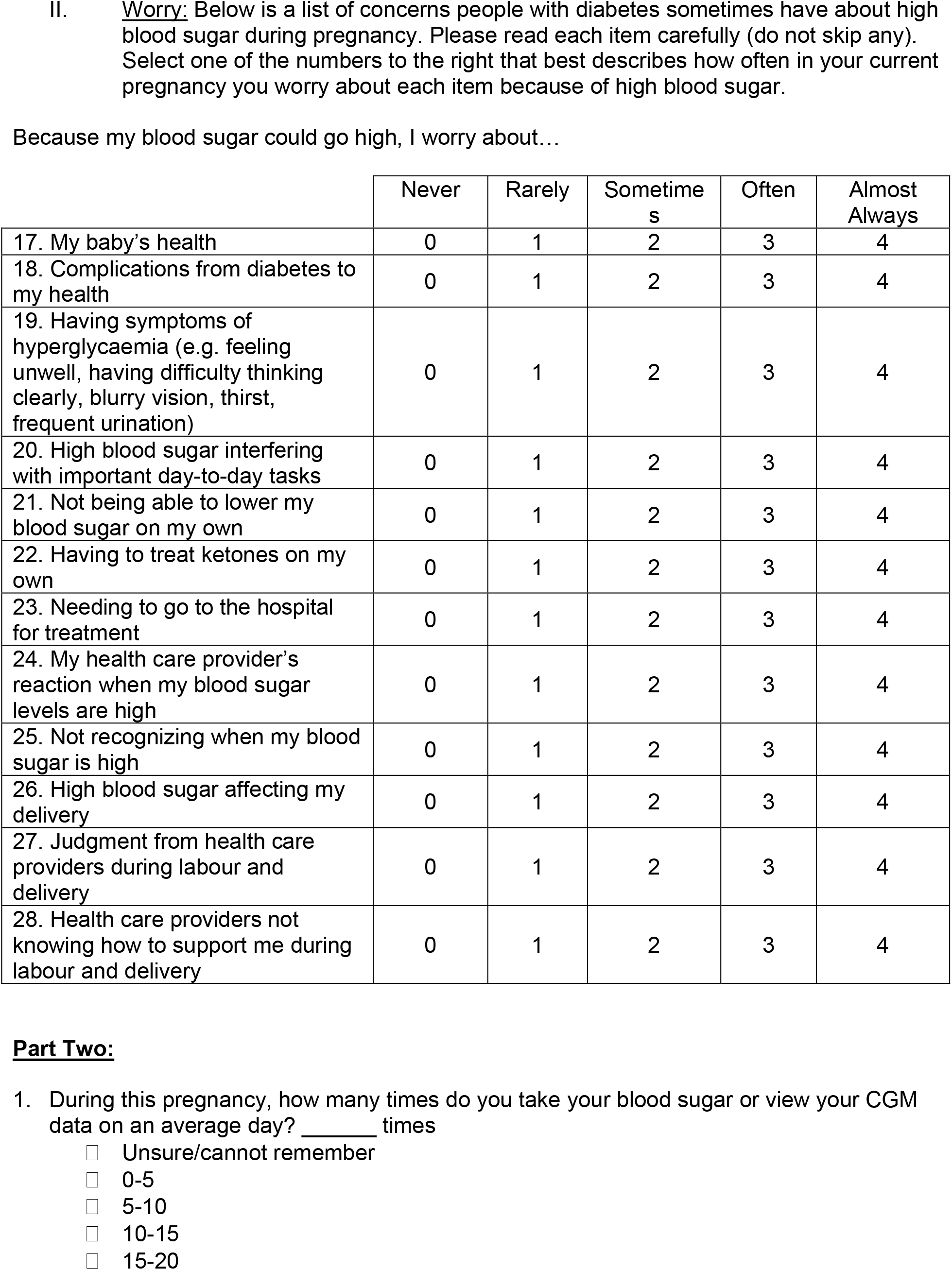
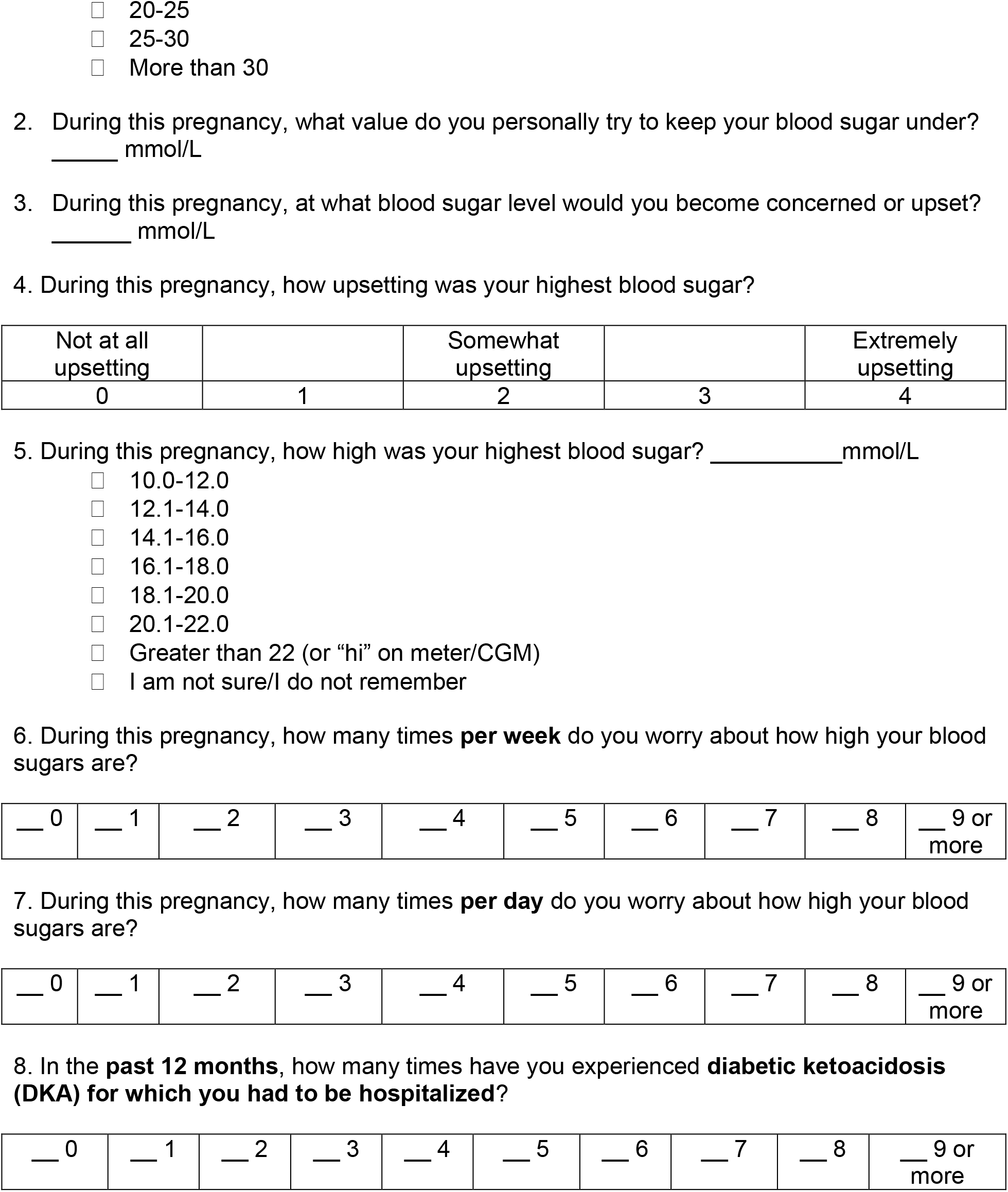
Fear of High Blood Sugar in Pregnancy Questionnaire.

|  | Never | Rarely | Sometimes | Often | Almost Always |
| --- | --- | --- | --- | --- | --- |
| 1. Ate smaller snacks/meals | 0 | 1 | 2 | 3 | 4 |
| 2. Delayed snacks/meals | 0 | 1 | 2 | 3 | 4 |
| 3. Skipped snacks | 0 | 1 | 2 | 3 | 4 |
| 4. Skipped meals | 0 | 1 | 2 | 3 | 4 |
| 5. Gave myself more insulin than I calculated or the pump-recommended for a correction bolus when my blood sugar was high (i.e. rage bolused) | 0 | 1 | 2 | 3 | 4 |
| 6. Reduced my out-of-town recreational travel | 0 | 1 | 2 | 3 | 4 |
| 7. Had someone else drive instead of me, when possible | 0 | 1 | 2 | 3 | 4 |
| 8. Decreased the amount of time I spent socializing with friends | 0 | 1 | 2 | 3 | 4 |
| 9. Avoided restaurants/food I did not prepare myself | 0 | 1 | 2 | 3 | 4 |
| 10. Stayed at home more than is typical for me | 0 | 1 | 2 | 3 | 4 |
| 11. Reduced my planned exercise/physical activity | 0 | 1 | 2 | 3 | 4 |
| 12. Added unplanned exercise to lower blood sugar | 0 | 1 | 2 | 3 | 4 |
| 13. Tried to avoid situations or tasks during which I would normally keep my blood sugar higher on purpose (e.g. work presentations) | 0 | 1 | 2 | 3 | 4 |
| 14. Tried to avoid stressors | 0 | 1 | 2 | 3 | 4 |
| 15. Stopped working/took leave from work | 0 | 1 | 2 | 3 | 4 |
| 16. Changed my job duties | 0 | 1 | 2 | 3 | 4 |
| 17. My baby's health | 0 | 1 | 2 | 3 | 4 |
| 18. Complications from diabetes to my health | 0 | 1 | 2 | 3 | 4 |
| 19. Having symptoms of hyperglycaemia (e.g. feeling unwell, having difficulty thinking clearly, blurry vision, thirst, frequent urination) | 0 | 1 | 2 | 3 | 4 |
| 20. High blood sugar interfering with important day-to-day tasks | 0 | 1 | 2 | 3 | 4 |
| 21. Not being able to lower my blood sugar on my own | 0 | 1 | 2 | 3 | 4 |
| 22. Having to treat ketones on my own | 0 | 1 | 2 | 3 | 4 |
| 23. Needing to go to the hospital for treatment | 0 | 1 | 2 | 3 | 4 |
| 24. My health care provider's reaction when my blood sugar levels are high | 0 | 1 | 2 | 3 | 4 |
| 25. Not recognizing when my blood sugar is high | 0 | 1 | 2 | 3 | 4 |
| 26. High blood sugar affecting my delivery | 0 | 1 | 2 | 3 | 4 |
| 27. Judgment from health care providers during labour and delivery | 0 | 1 | 2 | 3 | 4 |
| 28. Health care providers not knowing how to support me during labour and delivery | 0 | 1 | 2 | 3 | 4 |

|  |  |  |  |  |
| --- | --- | --- | --- | --- |
| Not at all<br>upsetting |  | Somewhat<br>upsetting |  | Extremely<br>upsetting |
| 0 | 1 | 2 | 3 | 4 |

|  |  |  |  |  |  |  |  |  |  |
| --- | --- | --- | --- | --- | --- | --- | --- | --- | --- |
| __ 0 | __ 1 | __ 2 | __ 3 | __ 4 | __ 5 | __ 6 | __ 7 | __ 8 | __ 9 or more |

7. During this pregnancy, how many times **per day** do you worry about how high your blood sugars are?
|  |  |  |  |  |  |  |  |  |  |
| --- | --- | --- | --- | --- | --- | --- | --- | --- | --- |
| __ 0 | __ 1 | __ 2 | __ 3 | __ 4 | __ 5 | __ 6 | __ 7 | __ 8 | __ 9 or more |

|  |  |  |  |  |  |  |  |  |  |
| --- | --- | --- | --- | --- | --- | --- | --- | --- | --- |
| __ 0 | __ 1 | __ 2 | __ 3 | __ 4 | __ 5 | __ 6 | __ 7 | __ 8 | __ 9 or more |

## Discussion

Diabetes management during pregnancy is demanding. Given the substantial role that self-management plays in diabetes care, assessment of evolving diabetes management strategies should evaluate person-reported outcomes of concern to those living with diabetes and responsible for day-to-day management, in addition to glycaemic and other medical outcomes. Studies of automated insulin delivery in pregnancy have demonstrated benefits in terms of decreased worry about glucose control and decreased burden of diabetes management in qualitative interviews.^5,12^ Qualitative interviewing, however, is costly, time-consuming, and can contribute to the burden of research participation during pregnancy, a period when diabetes management is already overwhelming.^13,14^ In addition, qualitative studies do not allow for the generation of quantitative evidence and are challenging to standardise across research practice, which limit the comparability of findings across different studies.

Person-reported outcome measures that are simple to administer and do not add substantially to the demands of research participation would be a preferred method of assessing the impact of evolving diabetes management strategies on quality-of-life outcomes. Most type 1 diabetes pregnancy trials to date have not shown benefits in the scores of commonly used PROMs,^2,7,12,15,16^ likely because the PROMs used do not effectively capture the outcomes of greatest importance to pregnant women with type 1 diabetes.^4^

Fear of hyperglycaemia is an important person-reported outcome among pregnant women with type 1 diabetes.^4^ Our questionnaire, developed using input from women with experience of type 1 diabetes and pregnancy, systematically quantifies fear of hyperglycaemia in pregnancy.

To our knowledge, this is the first questionnaire to evaluate fear of hyperglycaemia in pregnant women with type 1 diabetes. Polonsky et al. found that in a sample of adults with type 1 diabetes, asking three items related to hyperglycaemia aversiveness identified a subset of the sample with significantly greater time in range, lower glucose management indicator, and more severe hypoglycaemia.^17^ The motivation of those with hyperglycaemia aversiveness was thought to be avoidance of long-term complications of diabetes.^17^ Singh et al. developed the Hyperglycaemia Avoidance Scale, a 22-item questionnaire that quantifies preference for low blood glucose and worries and behaviours related to hyperglycaemia.^18^ These previous works are not specific to pregnancy, a time during which women with type 1 diabetes have increased fear of high blood sugar due to its potential consequences to their babies, rather than to themselves.^5,6,13^

A recent consensus statement emphasized the need to assess PROMs both in research and in clinical practice, since psychosocial functioning and mental health status profoundly influence self-management behaviours and overall health outcomes in diabetes.^19^ Listening to the perspectives of people with diabetes is important in determining which person-reported outcomes to assess, since their concerns and priorities may differ from the traditional clinical and research outcomes of glycaemic and medical complications. The Fear of High Blood Sugar in Pregnancy Questionnaire was created in response to our previous qualitative work, where we found that fear of hyperglycaemia is a prevalent concern amongst women with type 1 diabetes in pregnancy, and there is need for a PROM that evaluates this.^4^ A strength of our questionnaire is that it was developed by a member of the research team with lived experience of type 1 diabetes in pregnancy, and further revisions were made in collaboration with women with type 1 diabetes and pregnancy experience. Study participants had experience with various methods of insulin delivery during pregnancy; some had experienced multiple methods of insulin delivery across different pregnancies, including automated insulin delivery.

A limitation of our questionnaire is the lack of available data to assess its internal consistency, external consistency, reliability, and validity. We did not have access to participants’ clinical data to determine if scores on our questionnaire correlated to measures such as time in range, time below range, or incidence of severe hypoglycaemia. However, the person-oriented methods used in developing our questionnaire ensure that it has strong face and content validity as a PROM. Given the need for such a questionnaire in current diabetes in pregnancy research, we hope that this questionnaire will prove useful to researchers and that its broadened use will allow for future work to determine reliability and validity. Another limitation was that our study participants were of high socioeconomic status and the majority were white or Caucasian, which does not reflect the diversity of women with type 1 diabetes. Future work with this tool in a more diverse group of women with type 1 diabetes is needed.

Future studies should confirm the validity and reliability of this questionnaire in both research and clinical settings.

## Conclusions

Type 1 diabetes management is particularly challenging in pregnancy, and one source of stress for pregnant women is the fear of hyperglycaemia and its effects on their unborn children. This project created the Fear of High Blood Sugar in Pregnancy Questionnaire, which is the first step in the development of a pregnancy-specific PROM to be used to assess the quality-of-life impact of evolving diabetes management strategies for pregnancy.

## Data Availability

All data produced in the present work are contained in the manuscript

## Author Contributions

AD developed the first draft of the person-reported outcome measure. JG and AD interviewed participants. AD, KHC, and LED designed the study, and reviewed, edited, and approved the manuscript.

## Acknowledgements

The authors thank the participants of this study for sharing their experiences of type 1 diabetes in pregnancy. The authors thank L Gonder-Frederick and D Cox, creators of the Hypoglycaemia Fear Survey-II, whose work is the foundation for the questionnaire presented here. We thank J Booth for assistance with manuscript preparation.

## Notes

**Conflicts of Interest:** LED reports in-kind donations and reduced cost for study supplies for investigator-initiated trials from Medtronic, Dexcom, Tandem Diabetes Care, and Inter-analytics.

### Competing Interest Statement

Dr Donovan received in-kind donations and reduced cost for study supplies for investigator-initiated trials from Medtronic, Dexcom, Tandem Diabetes Care and Inter-analytics and speaker honoraria from Dexcom. Amy Dunlop reported receipt of personal fees from Tandem Diabetes Care for pump trainings and presentations on insulin pump therapy.

### Funding Statement

This study did not receive any funding

### Author Declarations

The Conjoint Health Research Ethics Board at the University of Calgary approved this study (REB 21-0447).

